# Impact of digital assistive technologies on the quality of life for people with dementia: A scoping review

**DOI:** 10.1101/2023.10.02.23296434

**Authors:** Charlotte Schneider, Marcia Nißen, Tobias Kowatsch, Rasita Vinay

## Abstract

**Background:** Digital assistive technologies (DATs) have emerged as promising tools to support the daily life of people with dementia (PWD) by offering support in various aspects of daily life. Quality of life (QOL) is an important consideration when discussing the care of PWD in relation to their autonomy. Current research tends to concentrate either on specific categories of DATs, or provide a generic view. Therefore, it would be of essence to provide a review of the different kinds of DATs, and how they contribute to improving QOL for PWD.

**Objective:** This scoping review aimed to review DATs and their impact on QOL for PWD.

**Method:** For this scoping review a broad literature search was performed in Cochrane, Embase, PubMed, Scopus, and Web of Science, covering scientific literature from January 2013 and May 2023. Screening and data extraction were conducted, followed by quantitative and qualitative analysis using thematic analysis principles and Digital Therapeutics (DTx) Alliance categories for DAT grouping.

**Results:** The literature search identified 6’083 records, with 1’056 duplicates. After screening, 4’560 full-texts were excluded, yielding 122 studies of different designs. The DATs were categorized into *digital therapeutics* (n=109), *patient monitoring* (n=30), *digital diagnostics* (n=2), *care support* (n=2), and *health system clinical software (*n=1). These categories were identified to impact various aspects of QOL: preserving autonomy, engagement, and social interaction, health monitoring and promotion, improving activities of daily living, improving cognition, maintaining dignity, managing behavioral and psychological symptoms of dementia (BPSD), and safety/surveillance.

**Conclusions:** Various DATs offer extensive support, elevating the QOL of PWD. *Digital therapeutics* are predominantly used for aging-in-place and independent living through assistance with daily tasks. Future research should focus on less-represented DHT categories, such as *care support, health & wellness* or *software* solutions. Observing ongoing DAT developments and their long-term effects on QOL remains essential.

**Strengths and limitations of this study:**

- The study conducted an extensive search across five electronic databases spanning a decade to identify relevant literature on DATs and their impact on the QOL for PWD.
- By excluding conference proceedings, book chapters, pilot, and feasibility studies, the review might have missed ongoing or planned research that could offer insights into different DATs or QOL impacts.
- While the scoping review approach allowed for a broad overview, it didn’t assess the quality of included studies or intervention effectiveness, potentially introducing bias and limiting in-depth analysis.
- The study’s emphasis on patient-facing DATs could have introduced bias, highlighting *digital therapeutics* in the included literature while potentially overlooking other assistive technology categories.

## Introduction

### Background

In 2023, more than 55 million individuals worldwide are affected by dementia, with approximately 10 million new cases diagnosed yearly [1]. Dementia encompasses various impairments regarding language, memory, cognition, and the ability to perform daily activities [1]. It progressively worsens over time and primarily affects individuals over 65, however can also effect those younger than 65, known as young onset dementia [1]. Globally, dementia currently ranks as the seventh leading cause of death, significantly contributing to disability and dependency among the older population [1]. The changing demographic landscape presents difficulties for caregivers and our healthcare system. As a result, there’s a growing focus on using digital assistive technologies (DATs) to address these challenges and help sustain the independence of people with dementia (PWD) [2].

DAT is an umbrella term covering technologies used for education and rehabilitation, to overcome participation and activity restrictions, and to improve cognitive, sensory, and motor abilities. It encompasses any technology that empowers individuals with functional constraints in everyday routines, educational pursuits, occupational endeavors, or recreational engagements [3]. DATs offer a valuable means for individuals and caregivers to manage various aspects of their daily routines effectively. They hold great promise for the care and support of PWD, and to alleviate the challenges associated with caregiving [4]. Recent technological advancements have paved the way for creating devices and applications that leverage sensory data tailored specifically for PWD. Notably, smartphones and wearables are now being employed to track physical activities, enabling in-home care assistance [4] and serving as location trackers for monitoring wandering behavior [5]. Moreover, the emergence of artificial intelligence (AI) has led to the development of social assistive robots. These robots are designed to provide companionship and engage in therapeutic activities, such as the robotic seal Paro, which serves as an illustrative example [6]. These technologies extend beyond basic assistance with daily tasks; they also contribute to the preservation of social interactions, memory support, participation in leisure activities, location tracking, and health monitoring [2][7].

Maintaining a good quality of life (QOL) is essential for PWD and must be considered when assessing the impact of DATs. QOL encompasses physical and mental well-being and extends to social and emotional dimensions (e.g., emotional stability, social integration, or self-esteem) [8]. Various tools such as questionnaires and self-assessment scales measure the overall perceived QOL; activity-based assessments or cognitive status evaluations serve as diverse means for quantifying QOL [9]. Consequently, these measures and evaluation instruments should be regarded as guiding tools in determining the QOL experienced by PWD.

While the current literature focuses on specific classes of DATs, often highlighting particular outcomes, there is a lack of a comprehensive overview of the different DATs and their impact on the QOL of PWD. This scoping review aims to fill this gap.

### Objectives

This scoping review aimed to provide an overview of DATs for PWD, and the ways in which they influence individual’s QOL. Therefore the following research question was formulated: “What is the impact of DATs on the QOL for people with dementia?”

In this scoping review, the opportunities of DATs and their role in enhancing caregiving practices and the QOL of PWD were investigated. Through a compilation of prevailing literature, this review can enhance the decision-making process by facilitating stakeholders’ comprehension of the spectrum of accessible DATs and their efficacy in effectively elevating the QOL as a primary objective.

## Methods

### Search Strategy

The methodological framework proposed by Arksey and O’Malley [10] and the recommendations from *PRISMA Extension for Scoping Reviews (PRISMA-ScR): Checklist and Explanation*[11] were adopted to conduct this scoping review. A protocol [12] was drafted and revised by authors CS, TK and RV. It was preregistered on the Open Science Framework Registry on 5 May 2023 (https://osf.io/zcnx8/).

Search terms were derived from a preliminary search and analyzed by comparing the words found in titles, abstracts, and keywords, following a ‘patient/population, intervention, outcome’ (PIO) concept. Additionally, to enhance the accuracy and comprehensiveness of the search results, all authors were involved in a consensus process, and an additional expert was consulted to validate the identified terms and suggest any additional relevant keywords. A comprehensive search was performed on 17 May 2023 across five electronic databases: Cochrane, Embase, PubMed, Scopus, and Web of Science, to locate published literature surrounding the research question. To focus on recent technological advancements, only articles published between 1 January 2013 and 17 May 2023 were considered, allowing for a more up-to-date review. The search terms using the PIO table and full electronic search strategy can be seen in Supplementary file 1.

### Inclusion and Exclusion Criteria

The articles were screened following specific inclusion and exclusion criteria, established by all authors and consolidated by an additional expert. Table shows the eligibility criteria to ensure the relevance of the included studies to the research question.

**Table 1.** Inclusion and Exclusion Criteria.

| Inclusion Criteria | Exclusion Criteria |
| --- | --- |
| Scientific literature (completed)<br>Articles that have a primary focus on DATs for PWD<br>Articles discussing perspectives of caregivers, family members, or healthcare workers in relation to a PWD<br>Articles about people living in diverse settings including communities, hospitals or nursing homes, and all severities of dementia. We will not use age as a criterion. | Articles not in English or German<br>Articles that discuss dementia negligibly or with other comorbidities/health conditions<br>Articles that mention DATs briefly or as an insignificant part of a review<br>Pilot or feasibility studies that only report the implementation of an intervention<br>Book chapters, commentaries, conference proceedings, editorials, interviews, opinion pieces, proposals, reports, protocols, short news<br>Non-human studies |

### Screening, Data Extraction, and Analysis

The search results were extracted and uploaded onto a literature review software, Rayyan (www.rayyan.ai) for screening. From May to June 2023, authors CS and RV screened all eligible articles’ titles and abstracts to determine their suitability for a full-text review according to the inclusion and exclusion criteria. Potential discrepancies were discussed between the authors and resolved through discussion and consensus. Author CS piloted the data extraction using five articles, checked and consulted by RV. An example of the data extraction form is described in **Error! Reference source not found.** A full-text review of the final sample of included studies was conducted by CS and further consolidated by RV. A critical appraisal of individual sources of evidence was not done for this scoping review.

A narrative synthesis accompanied with frequency analysis was performed of the included literature to present findings on (1) author locations, (2) study approach (3) type of article (4) study locations, (5) digital health technology (DHT) categories (see Coding Strategy), (6) target population, and (7) instruments measuring QOL. The QOL measures were initially evaluated according to the protocol [12]. Based on a pilot of the extraction of the articles, it became obvious that there was rarely one distinct “QOL instrument” used throughout all articles. However, these metrics were discovered to be excessively diverse and unsuitable to be reported consistently, leading to their omission. Instead, indirect outcomes, which also influence the QOL, were additionally recorded (e.g., activity instruments, cognitive status, rating of the individual’s QOL, etc. [9]). Furthermore, the protocol outlined the analysis of the type of DATs and sensory distribution channels [12]. During the extraction of the articles, it became evident that the type of DATs and sensory distribution channels wouldn’t provide consistent reporting across the studies, and were instead replaced with DHT categories. Several studies discussed different DATs employing distinct sensory distribution channels, making it impossible to provide a uniform report, resulting in their exclusion.

### Coding strategy

The codes for QOL measures/outcomes were generated by authors CS and RV based on thematic analysis [13]. The *Flanagan Quality of Life Scale* served as the basis for the code, which forms the general ideas about concepts for evaluating the QOL [16]. In addition, QOL scales such as QOL-AD, and DQOL were explored to identify relevant concepts for PWD [17]. Physical, social, and environmental concepts were also included to have a broader impact on QOL than just health-related QOL [18]. Ethical considerations were integrated into the coding strategy by drawing upon principles of biomedical ethics (i.e., dimensions such as autonomy, and dignity) [19]. Furthermore, it should be noted that the priorities of these principles may shift as an individual’s dementia progresses. For instance, autonomy might be more important for individuals with mild dementia, while maintaining dignity becomes paramount for those with severe dementia. This consideration enabled providing a holistic overview of QOL dimensions across all stages of dementia, aligning with the evolving ethical dimensions of care. CS applied the codes to the articles using the ATLAS application (www.atlasti.com), and uncertainties were resolved by RV.

All the included studies were initially categorized using an inductively created classification scheme. However, it became apparent that this method was inadequate for representation purposes. Subsequently, the classification scheme was deductively realigned with the Digital Therapeutics (DTx) Alliance categories, and all the studies were reclassified accordingly.

The DTx Alliance introduced eight (industry- and admin-, healthcare provider, and patient-facing) DHT categories: non-health system software/ digital health solutions, health system operational software, health system clinical software, health & wellness, patient monitoring, care support, digital diagnostics, and digital therapeutics [14]. To apply these categories to the included studies, some of the definitions were adapted (see

Table for an adapted version of the definitions):

- Initially, the *patient monitoring* category exclusively monitored specific patient health data. However, in the context of this scoping review, location data was included as well to be able to classify studies that discuss tracking devices.
- Only studies explicitly mentioning self-management or similar concepts were assigned to the c*are support* category. Otherwise, nearly all papers would qualify for this category, as they predominantly aim to assist patients in various ways.
- The criteria for the *digital therapeutics* category have been refined and a broader range of digital technologies was encompassed that generate and deliver medical interventions, while the initial definition pertained to “health software designed to treat or alleviate a specific disease or medical condition by generating and delivering a medical intervention” [14]. It is important to note that not all of them qualify as medical devices or products, and may not be subject to regulation based on the adapted definition for the purpose of this scoping review.

**Table 2.** The eight DHT categories with their adapted definitions and examples. Source: DTx Alliance (https://dtxalliance.org) [14], [15].

|  |  |  |
| --- | --- | --- |
| Industry and admin-facing | Healthcare providers - facing | Patient-facing |

| DHT Category | Non-health system software/DH solutions | Health system operational software | Health system clinical software | Health & wellness | Patient monitoring | Care support | Digital diagnostics | Digital therapeutics |
| --- | --- | --- | --- | --- | --- | --- | --- | --- |
| **Overview** | Health Information Technology (HIT) and digital health (DH) solutions designed for stakeholders outside of the hospital and health system settings, such as pharma, medtech, insurances, employers and pharmacies.) | Enterprise HIT aimed at delivering benefits and facilitating support for non-clinical functions ( i.e., operational and financial aspects). | Enterprise HIT and DH solutions designed to offer clinicians assistance in effectively managing their patient populations.<br>Divided into four subcategories: clinical documentation & image archiving, communication support, clinical decisions support, and telehealth [15] | DH solutions that are not specific to any particular disease and primarily focus on capturing and storing general health information while encouraging and facilitating a healthy lifestyle , and do not deliver any medical interventions. | Digital solutions to monitor specific (health or location) data that can be interpreted by a physician to assist in decision making for a medical procedure, or informs a caregiver or family member. | Digital solutions created to assist patients in improving their self-management of a specific disease or medical condition. May offer recommendations, or information s but they do not deliver medical treatments. | Verified digital tools and software that provide a diagnosis or prognosis for a particular disease or medical condition. | Any digital technology that address or mitigate a disease by generating and implementing a medical intervention. |
| **Examples** | Accounting Software, documentation tool for compliance. | Software for patient bed management, data & analytics platforms. | Platform for electronic medical records and clinical communications. | Motivational tools, health diaries. | Tracking devices detecting falls, GPS, wearables. | Static care support (medication tracker application) , responsive care support. | Apps for dementia screening, algorithm returning a measure of Alzheimer's disease severity. | Robots, Apps, multimedia systems, Augmented Reality/ Virtual Reality. |

## Results

### Search Findings

The search identified 6’083 records from the electronic databases. A total of 1’056 duplicates were identified and removed, leaving 5’027 articles and trial records to be screened. Title and abstract screening led to the exclusion of 4’560 records, resulting in 467 full texts that needed to be assessed for eligibility. Of these, 122 studies were included (see <u>Supplementary file 2</u>), and 345 were excluded for the following reasons: 72 did not focus only on PWD or their results could not be separated (wrong population), 69 had the wrong design, and 59 pursued wrong aims or outcomes, 42 were the wrong publication type or language (i.e., editorials, book chapters, protocols, news articles, non-English), 30 were only trial registry records, 26 could not be accessed, 25 were either pilot, feasibility or usability studies which only tested the viability of intervention, 12 did not deal with QOL, and 10 did not include DATs. These are reported in the PRISMA flowchart in Figure 1.

**Figure 1.**
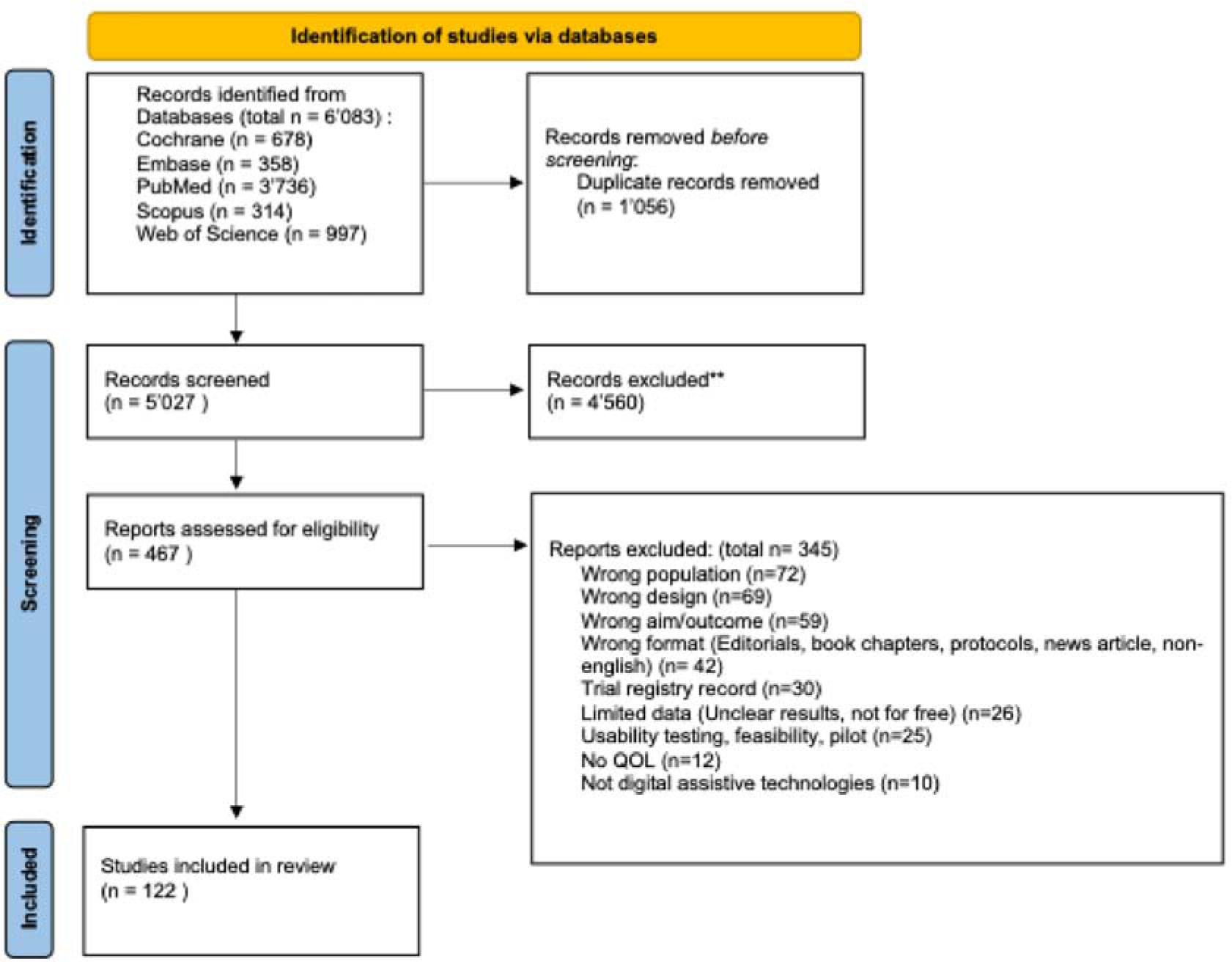
PRISMA flow chart.

### Characteristics of Included Studies

Scientific literature from 1 January 2013 to 17 May 2023 was included to capture the most recent findings. Figure 2 represents the number of articles published for each year in the date range included for this scoping review.

**Figure 2.**
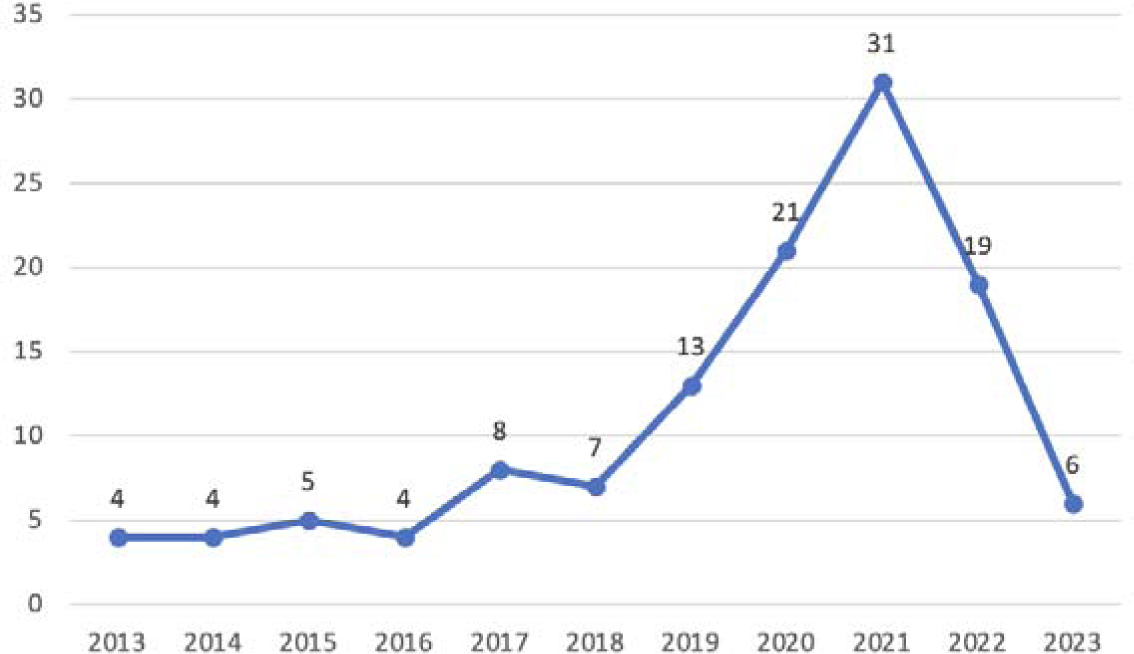
The graph displays the trend of the number of studies published per year.

The authors’ location was collected for each study, ensuring unique country attribution per study while capturing all countries affiliated with each author. There were 180 entries recorded. The countries with the highest number of entries were the United Kingdom (n=38, 21.1%), Australia (n=17, 9.4%), the United States (n=14, 7.8%), Canada, China, and Norway (n=11, 6.1% each). Notably, European countries showed significant numbers of contributions, including Germany and the Netherlands (n=8, 4.4% each). Sweden and Italy (n=6, 3.3% each) and France (n=4, 2.2%). From the Asian and Oceania continent, contributions were made by Japan (n=6, 3.3% each), Taiwan and South Korea (n=5, 2.8% each), New Zealand and Singapore (n=3, 1.7% each), Pakistan and Qatar (n=2, 1.1% each), and India, Indonesia, and Malaysia (n=1, 0.6% each). From the African continent, there were only contributions from South Africa (n=4, 2.2%). Some countries, such as Brazil, Cyprus, Czech Republic, Denmark, Greece, Malta, Portugal, Spain, and Switzerland, had limited representation, with only 1-2 (0.6% respectively 1.1%) contributions each (Figure 3).

**Figure 3.**
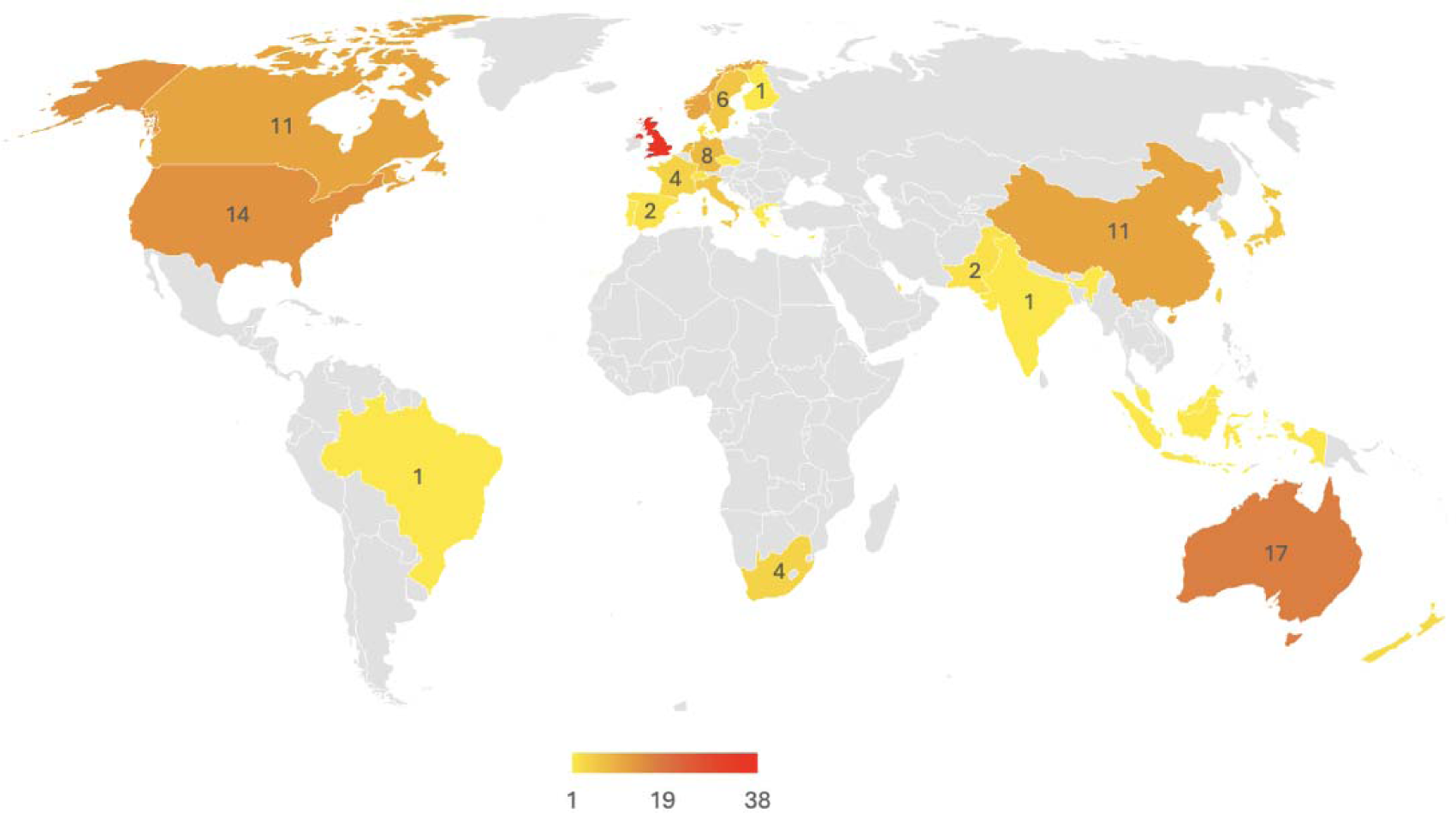
Distribution of the authors’ locations. The countries are colored based on the number of country attribution, while encompassing all countries associated with each author.

A variety of study designs were used in the included studies (Figure 4), comprising of 56 (46%) reviews, 16 RCTs (13%), 15 (12%) case studies, 13 (11 %) intervention studies, 8 (6%) pilot or feasibility studies, 5 (4%) interviews/surveys, 5 (4%) trials, 2 (2%) cohort studies, and 2 (2%) observation studies.

**Figure 4.**
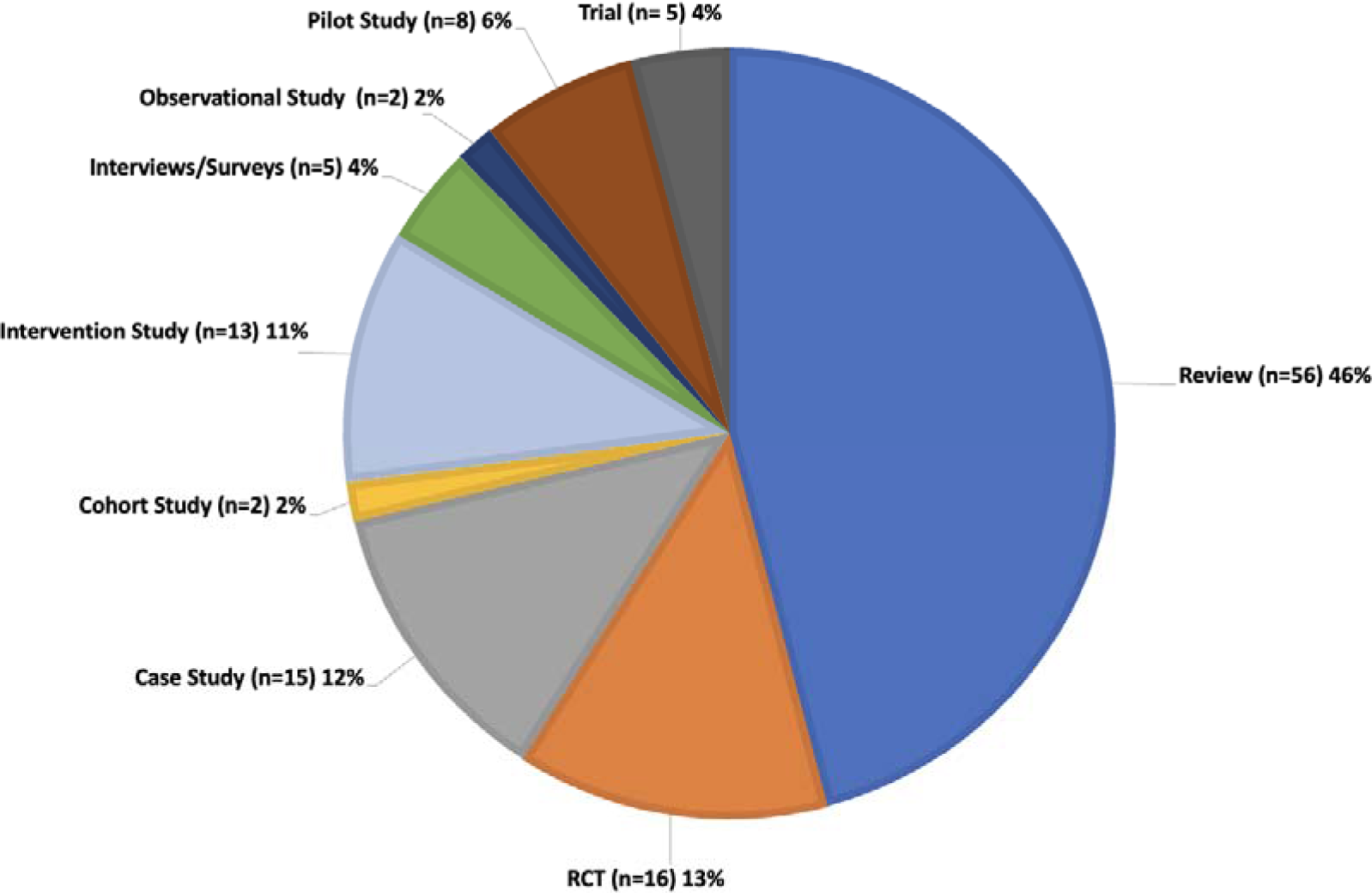
The distribution of the study designs.

In total, there were 36 (30%) quantitative studies, 52 (43%) qualitative studies, and 34 (28%) adopted a mixed-methods approach.

The study locations were diverse with 73 different study locations identified. The highest number of studies were carried out in the UK (n=15, 20.5%), followed by Australia (n=9, 12.3%), the USA (n=7, 9.6%), Sweden (n=6, 8.2%), and Norway (n=5, 6.8%). Other locations included Canada, Italy, and the Netherlands (n=4, 5.5% each), Denmark and France (n=3, 4.1% each), and Japan and Taiwan (n=2, 2.7% each). There was one study (1.4%) each from China, Cyprus, Germany, Ghana, Greece, Korea, New Zealand, Singapore, and Spain. Within these locations, different study settings were utilized; home-based (n=25, 33.8%), residential care (n=10, 13.5%), long-term care and nursing homes (n=9, 12.2% each), daycare centres (n=7, 9.5%), community-based and hospital (n=4, 5.4% each), other care facilities, or memory clinics (n=3, 4.1% each), and a laboratory setting (n=1, 1.4%).

Diverse target groups of the included studies were identified, as presented in Table. As indicated in the review process and as part of the exclusion criteria, only research articles that could separate results for PWD from other comorbidities or conditions were included.

**Table 3.**
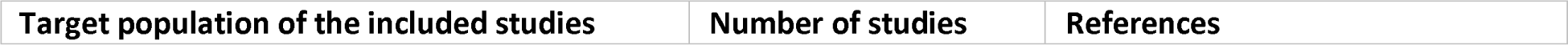
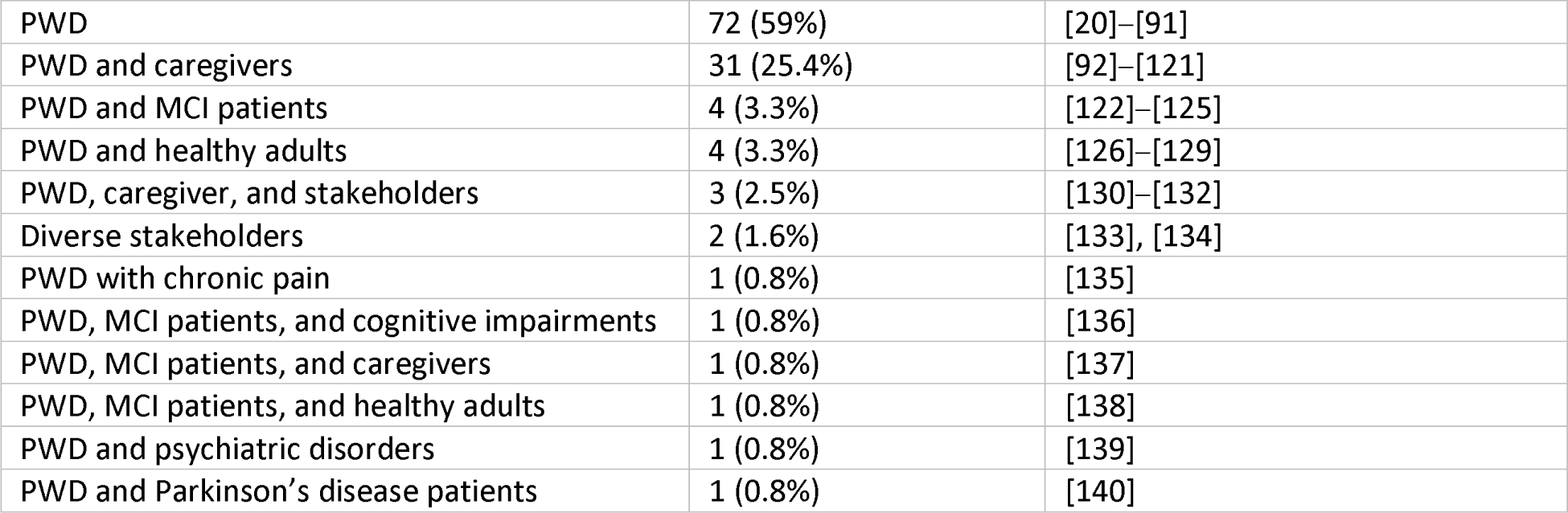
The different target groups of the included studies sorted by number of studies. The frequency of each class is indicated (as number of studies), along with the corresponding references.

### Characteristics of DATs

#### Categories of DATs

The DATs of the 122 included studies underwent classification into the DHT categories provided by the DTx Alliance [14]. Some studies discussed different DATs and therefore were assigned to multiple categories due to their varying applicability (see Table).

**Table 4.**
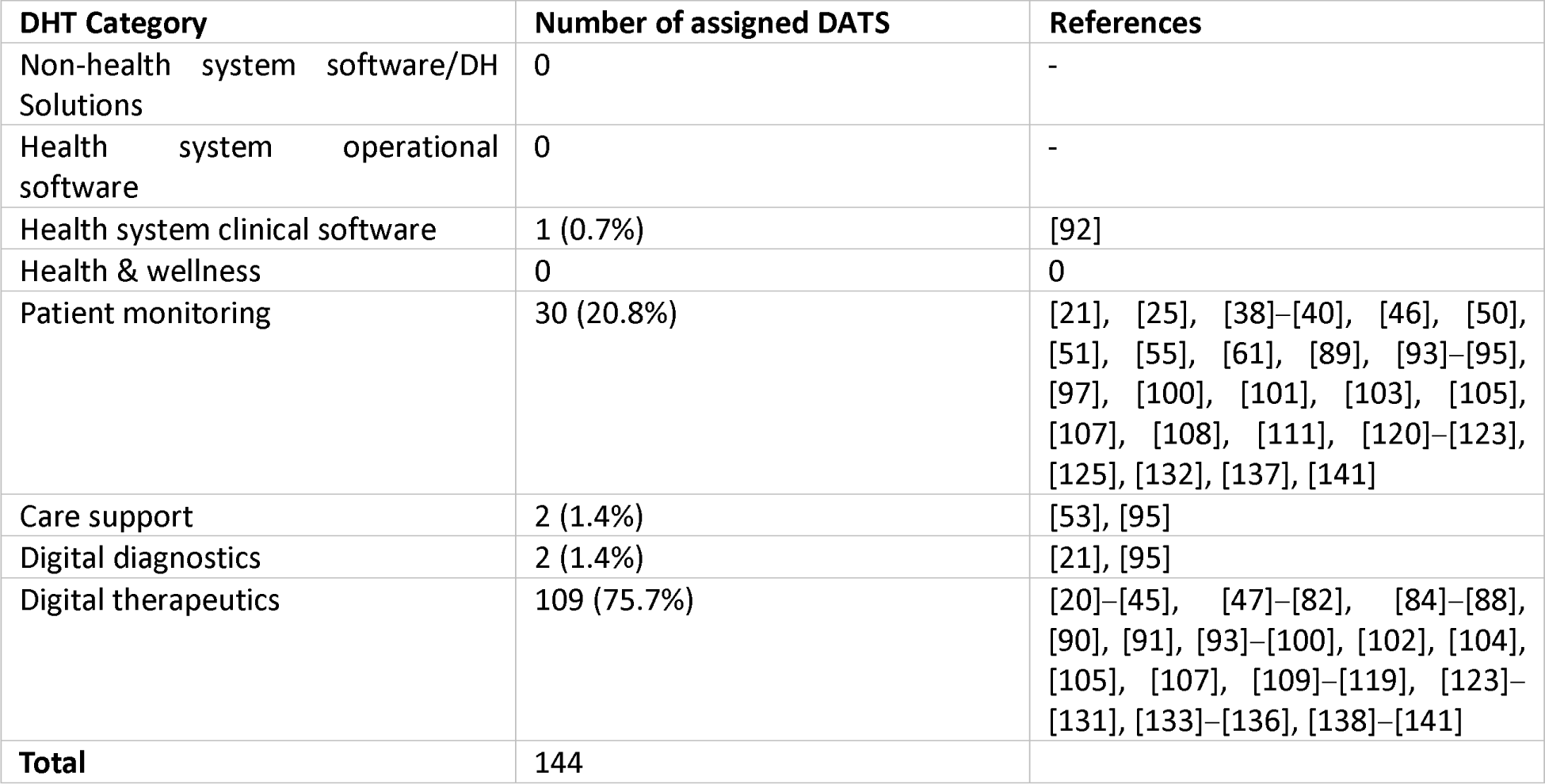
The different classes of DATs. The frequency of each category is indicated, along with the corresponding references.

#### Qualitative aspects of QOL

##### Positive impact of DATs on QOL dimensions

Through qualitative analysis of the included studies, recurring themes were identified that reflect the different aspects of how DATs influence QOL for PWD: preserving autonomy, engagement and social interaction, health monitoring and promotion, improving activities of daily living (ADL), improving cognition, maintaining dignity, managing behavioral and psychological symptoms of dementia (BPSD), and safety/ surveillance (Table 1). These concepts can influence each other, and it is possible for a single DAT to simultaneously have multiple effects. In the following sections, these impacts will be discussed in more detail based on the analysis of the included studies.

**Table 1.**
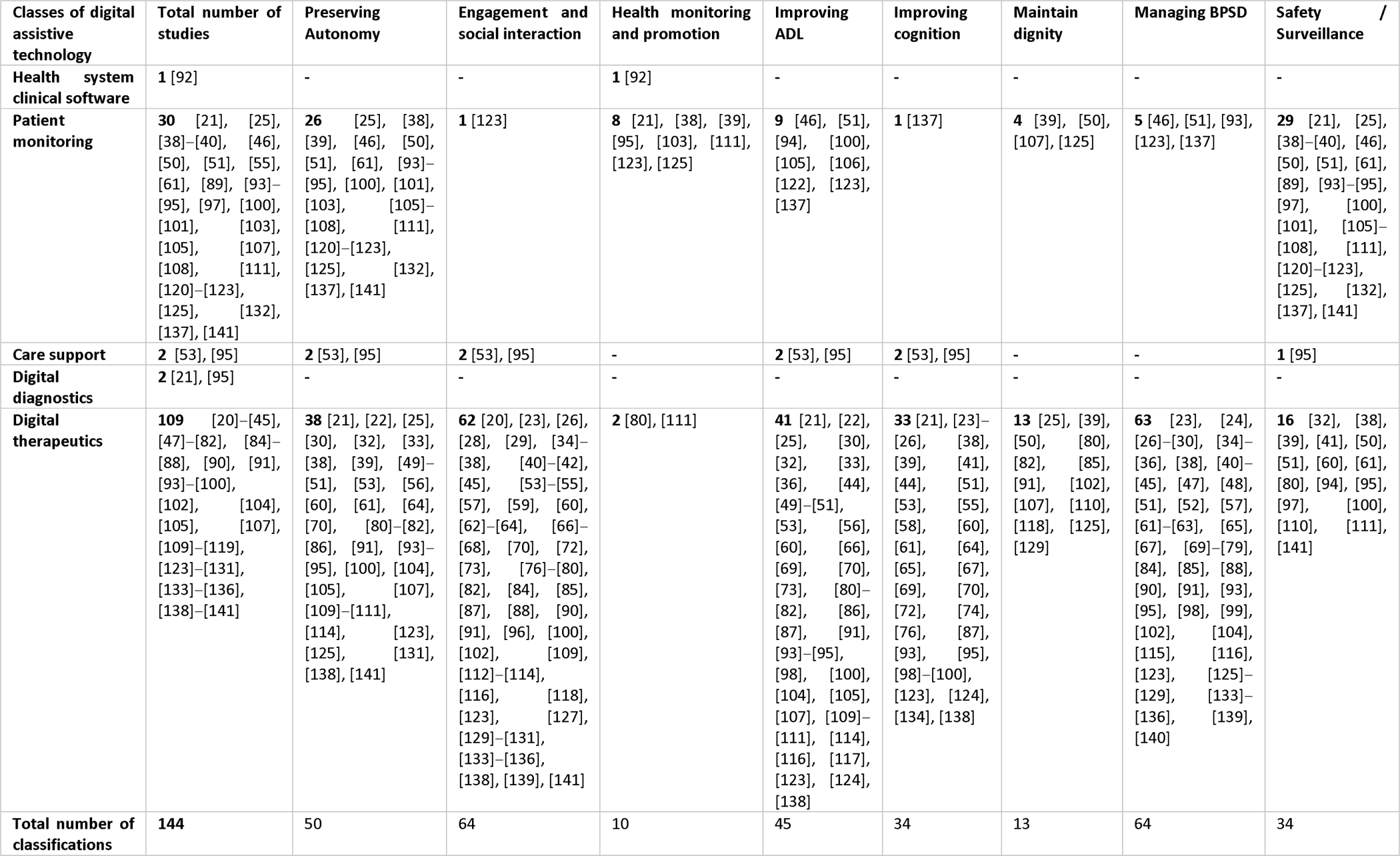
Overview of the different DHT categories with the various impacts on QOL and their frequency and references.

### Preserving Autonomy

Autonomy can be understood as an individual’s ability to be involved in decision-making, consent, receiving treatment or intervention (i.e., to choose and act independently without coercion) [19]. Within healthcare, the preservation of autonomy has emerged as a significant topic, most importantly known as the principle of respect for autonomy. DATs can be seen as a useful tool for promoting autonomy and, therefore, positively contributing to PWD’s lives. The use of DATs is intended to increase the capacity of an individual to participate in decision-making [122], [101] and promote independent living, and enable aging-in-place [39].

From 50 of the included studies, the following were used to promote autonomy: *patient monitoring* [25], [38], [39], [46], [50], [51], [61], [93]–[95], [100], [101], [103], [105]–[108], [111], [120]–[123], [125], [132], [137], [141], *care support* [53], [95] and *digital therapeutics* [21], [22], [25], [30], [32], [33], [38], [39], [49]–[51], [53], [56], [60], [61], [64], [70], [80]–[82], [86], [91], [93]–[95], [100], [104], [105], [107], [109]–[111], [114], [123], [125], [131], [138], [141].Resilience was also mentioned as a significant factor contributing to enhanced autonomy [41], [62], [129], [130].

### Engagement and social interaction

The topic of engagement and social interaction was highlighted in 64 studies. They differentiated between cases where DATs increased social interaction [20], [28], [29], [34], [35], [38], [41], [45], [54], [59], [64], [70], [72], [80], [82], [83], [85], [100], [102], [112], [113], [123], [129], [131], [133], [134] and fostered higher engagement (i.e., in therapy settings) [23], [26], [29], [34], [36]–[38], [40]–[42], [45], [53]–[55], [57], [60], [62], [63], [66]–[68], [70], [73], [76]–[79], [84], [85], [87], [88], [90], [91], [96], [100], [109], [112], [114], [116], [118], [122], [127], [130], [131], [133], [135], [136], [138], [139], [141].

62 of the included studies demonstrated that *digital therapeutics* play a significant role in promoting social interaction and engagement [20], [23], [26], [28], [29], [34]–[38], [40]–[42], [45], [53]–[55], [57], [59], [60], [62]–[64], [66]–[68], [70], [72], [73], [76]–[80], [82], [84], [85], [87], [88], [90], [91], [96], [100], [102], [109], [112]–[114], [116], [118], [123], [127], [129]–[131], [133]–[136], [138], [139], [141], including other DATs from the *patient monitoring* [123] and *care support categories* [53], [95].

### Health monitoring and promotion

Health monitoring refers to monitoring bodily functions and reporting their states or imbalances. 10 studies reported using DATs to monitor and promote health for PWD [21], [38], [39], [80], [92], [95], [103], [111], [123], [125]. This was achieved, for example, through social assistive robots [80] and wearables [103], including wristwatches that measure movement, skin temperature, and pulses [111].

### Improving activities of daily living (ADL)

DATs support PWD in maintaining or regaining their ability to perform activities of daily living (ADL) such as meal preparation, managing medication or communication. 45 of the included studies highlight this field’s broad spectrum of possibilities, for instance, they range from the implementation of interventions from the *patient monitoring* category [46], [51], [94], [100], [105], [106], [122], [123], [137], *care support* [53], [95], to *digital therapeutics* [21], [22], [25], [30], [32], [33], [36], [44], [49]–[51], [53], [56], [60], [66], [69], [70], [73], [80]–[82], [86], [87], [91], [93]–[95], [98], [100], [104], [105], [107], [109]–[111], [114], [116], [117], [123], [124], [138].

### Improving cognition

The varied application of DATs can enhance cognitive abilities, as indicated by 34 studies. They demonstrated that *patient monitoring* [137]*, care support* [53], [95]*, digital therapeutics* [21], [23]–[26], [38], [39], [41], [44], [51], [53], [55], [58], [60], [61], [64], [65], [67], [69], [70], [72], [74], [76], [87], [93], [95], [98]–[100], [123], [124], [134], [138] can lead to improvements in cognitive functioning.

### Maintaining dignity

In contrast to autonomy, dignity refers to the ability to preserve self-respect and personhood, while being recognized and valued in society, and discussed in 13 studies using diverse DAT categories: *digital therapeutics* [39], [50], [107], [125] and *patient monitoring* [25], [39], [50], [80], [82], [85], [91], [102], [107], [110], [118], [125], [129]. These categories are specifically designed to uphold and promote aging in place, as demonstrated in the review of Gettel et al. [39], by the use of pet robots to maintain the resident’s dignity and self-worth [102].

### Managing Behavioral and Psychological Symptoms of Dementia (BPSD)

There are 12 behavioral and psychological symptoms of dementia (BPSD), which include aberrant motor behavior, agitation, anxiety, apathy, appetite changes, delusions, depression, disinhibition, elation/euphoria, hallucinations, irritability, and sleep changes [142]. Different BPSD symptoms were shown to be managed by DATs in 64 studies through *digital therapeutics* [23], [24], [26]–[30], [34]–[36], [38], [40]–[45], [47], [48], [51], [52], [57], [61]–[63], [65], [67], [69]–[79], [84], [85], [88], [90], [91], [93], [95], [98], [99], [102], [104], [115], [116], [123], [125]–[129], [133]–[136], [139], [140] or by *patient monitoring* devices [30], [34], [36], [63], [76], [95], [115]. All the DATS were reported to have a positive effect on BPSD.

### Safety/Surveillance

34 of the included studies demonstrated that DATs can enhance the safety of PWD, for instance through *patient monitoring* [21], [25], [38]–[40], [46], [50], [51], [61], [89], [93]–[95], [97], [100], [101], [105]–[108], [111], [120]–[123], [125], [132], [137], [141], *care support* [95], and *digital therapeutics* [32], [38], [39], [41], [50], [51], [60], [61], [80], [94], [95], [97], [100], [110], [111], [141].

#### Negative impacts of DATs on QOL dimensions

Among the included studies, 18 studies reported negative impacts of DATs on the QOL [24], [43], [50], [52], [60], [70], [73], [79], [80], [88], [91], [104], [109], [114], [120], [125], [129], [138]. These negative impacts included increased anxiety [73], [91], [129], worsened agitation [43], concerns about negative consequences [70], confusion [60], worsened BPSD [43], [60], [100], increased hallucinations and decreased mood [60], anxiety towards the technology [72], [91], [129], as well as aggression, rejection or disliking the technology [129].

#### Instruments measuring the QOL

As demonstrated, DATs can diversely impact aspects of QOL. It was observed that numerous studies employ specific measures to assess these impacts, which can manifest in various forms. During the charting process, 33 studies revealed the utilization of different units of measurement in this context. In 18 studies, the *Quality of Life in Alzheimer’s Disease Scale (QOL-AD)* was utilized [25], [28], [30], [41], [43], [44], [52], [62], [65], [67], [70], [73], [85], [90], [91], [125], [130], [136], and in 11 studies the *Quality of Life in Late-Stage Dementia scale (QUALID)* [27], [35], [41], [43], [61], [73], [85], [91], [104], [126], [136]. Additionally, the *self-reported Dementia Quality of Life measure (DEMQoL)* was employed in 4 studies [73], [92], [104], [112], the *Dementia Quality of Life Instrument (DQoL)* in 3 studies [41], [61], [93], the *EQ-5D-5L* in 3 studies [99], [104], [110]. Other measuring instruments were used only once in each case: *Quality of Life (GQL8)* [124], *Quality of Life for People with Dementia (QUALIDEM*) [41], *Quality of Life Alzheimer’s disease (QoL) scale*[98], *SF-36 quality of life instrument* [103], *Cantril QoL ladder* [63], Carer-Qol-7D [112], *EUROHIS-QoL-8 and EuroQoL 5 Dimension Questionnaire* [91].

## Discussion

This scoping review identified 122 studies published between January 2013 and May 2023. The studies encompassed a range of diverse study designs, with the most prevalent category being reviews, constituting 56 of the total. This was followed by 16 RCTs and 15 case studies, representing the three most common study types. The origin countries of the authors exhibited a wide diversity, as did the locations where the studies were conducted. Among the identified studies, 72 focused on PWD as the target group. Subsequently, 30 studies concentrated on PWD and caregivers. The remaining studies explored various combinations, including examining and comparing PWD and other medical conditions or including other stakeholders in the analysis.

DATs were categorized into the *DTx Alliance* categories, with the largest category being *digital therapeutics* (n=109), followed by *patient monitoring* (n=30), *digital diagnostics* (n=2), *care support* (n=2), and *health system clinical software* (n=1). This distribution can be attributed to the search strategy, which specifically targeted DATs with a therapeutic focus.

This review highlights that DATs have the potential to impact the QOL of PWD in several identified thematic areas: preserving autonomy, engagement, social interaction, health monitoring and promotion, improving ADL, cognition, maintaining dignity, managing BPSD, and safety/surveillance. The different categories of DATs can provide diverse forms of support to PWD across these thematic areas, enhancing their overall QOL. DATs help PWD to age in place, live independently, and maintain their dignity, especially when supported by DATs that help PWD keep or regain their ability to engage in daily activities. DATs were also shown to help promote health by tracking bodily functions, and encouraging engagement and social contact. Overall, impacts could be seen on individual’s cognitive abilities, to manage BPSD, and improvements to their general well-being and safety.

However, in a few instances, negative impacts of DATs on the QOL of PWD were also identified. For example, worsened BPSD, worries about negative results, anxiety and aggression towards the technology, or disliking the technology.

In addition to the qualitative impacts, various quantitative quality-of-life instruments were examined. It was found that 12 different instruments were utilized across 18 of the included studies. The most frequently employed instrument was the *Quality of Life in Alzheimer’s Disease Scale (QOL-AD).* Furthermore, it was observed that DATs are utilized to support therapy [61], and can serve as therapeutic tools to even constitute a distinct form of therapy in themselves [23].

## Strengths and Limitations

This scoping review first attempted to provide an extensive overview of various DATs and their impact on the QOL for PWD. A comprehensive search strategy was implemented to achieve this, covering five electronic databases over a decade. However, considering the rapidly expanding landscape of DATs and the exclusion of conference proceedings, book chapters, pilot and feasibility studies, it is conceivable that this scoping review might overlook ongoing or planned studies that could shed light on alternative DATs or different impacts on QOL.

Conducting a scoping review instead of a systematic review has its limitations. In this approach, an assessment of the quality of the included studies or an evaluation of intervention effectiveness needs to be incorporated. Therefore the validity of interventions on their measured outcomes may present bias or need to be better analyzed and reported in included studies. The search strategy prioritized patient-facing DATs, thereby introducing a potential bias that elucidates the high predominance of *digital therapeutics* in the included literature.

Nonetheless, due to the restricted availability of evaluative research in this domain, the primary objective was to explore the impact of digital assistive technologies on the QOL for PWD.

## Conclusion

A variety of DATs are available, offering versatile applications and holding the potential to serve as promising instruments for enhancing the QOL among PWD. Further research must be conducted to examine the ongoing developments in DATs and their increasing impact on QOL, including their long-term effects, and a deeper conceptual understanding of how certain interventions correlate to improving QOL. Moreover, future research could consider placing a particular emphasis on the less-represented DHT categories, such as *care support, health & wellness* or *software*. Digital innovations offer significant potential in addressing the global increase in the elderly population and revolutionizing various aspects of elderly care [4]. Considering future research directions, for example in the context of voice assistants and advancements in large language models (LLM) [143], [144], would be of importance, as they facilitate the development of an interface that is more intuitive and natural, without demanding a high degree of dexterity.

## Supporting information

PRISMA-ScR Checklist

## Data Availability

All data produced in the present work are contained in the manuscript

## Author contributions

All authors have made substantial intellectual contributions to developing this scoping review and its revisions. The search question was conceptualized by TK and RV, and further developed by CS. The review approach and design were conceptualized by RV, with advice from TK. CS and RV developed and tested search terms with input and revisions from TK. CS and RV jointly screened all the studies resulting from the search strategy. CS conducted the full-text data extraction, with RV, MN, and TK reviewing and providing consultation. CS drafted the first manuscript, and all authors were involved in the revision of the manuscript. All authors approved the final version of the manuscript.

