## Supplementary file 1 for "Impact of digital assistive technologies on the quality of life for people with dementia: A scoping review"

Supplementary Table A. The search terms derived for the PIO framework.

| **Concept** | **Terms** |
| --- | --- |
| Population:  People with dementia | alzheimer* OR dement* OR early-onset OR frontotemporal lobar degeneration OR lewy-body dementia OR mixed dementias OR vascular dementia OR young onset |
| Intervention: Digital assistive technologies | digital assistive tool* OR digital assistive technolog* OR gerontechnolog* OR mobile OR robot* OR supportive technolog* OR technolog* assistive device* OR voice assistant* OR wearable device* OR wearable technolog* |
| Outcome: Quality of life | activities of daily living OR independence OR life quality OR living standards OR mental health OR perception OR physical health OR satisfaction OR quality of life OR qol OR safety OR standard of living OR value of life OR well-being |

*Supplementary Table B. The full electronic search strategy for all five databases; Cochrane, Embase, PubMed, Scopus, Web of Science.*

| **Date of Search** | **Electronic Database** | **Keyword searched** | **Number of studies** |
| --- | --- | --- | --- |
| 17.05.2023 10:56 | Cochrane | (alzheimer* OR dement* OR early-onset OR frontotemporal lobar degeneration OR lewy-body dementia OR mixed dementias OR vascular dementia OR young onset):ti,ab,kw AND (digital assistive tool* OR digital assistive technolog* OR gerontechnolog* OR mobile OR robot* OR supportive technolog* OR technolog* assistive device* OR voice assistant* OR wearable device* OR wearable technolog*):ti,ab,kw AND (activities of daily living OR independence OR life quality OR living standards OR mental health OR perception OR physical health OR satisfaction OR quality of life OR qol OR safety OR standard of living OR value of life OR well-being):ti,ab,kw | 678 |
| 17.05.2023  11:08 | Embase | (alzheimer*:ti,ab,kw OR dement*:ti,ab,kw OR 'early onset':ti,ab,kw OR 'frontotemporal lobar degeneration':ti,ab,kw OR 'lewy-body dementia':ti,ab,kw OR 'mixed dementias':ti,ab,kw OR 'vascular dementia':ti,ab,kw OR 'young onset':ti,ab,kw) AND ('digital assistive tool*':ti,ab,kw OR 'digital assistive technolog*':ti,ab,kw OR gerontechnolog*:ti,ab,kw OR mobile:ti,ab,kw OR robot*:ti,ab,kw OR 'supportive technolog*':ti,ab,kw OR 'technolog* assistive device*':ti,ab,kw OR 'voice assistant*':ti,ab,kw OR 'wearable device*':ti,ab,kw OR 'wearable technolog*':ti,ab,kw) AND ('activities of daily living':ti,ab,kw OR independence:ti,ab,kw OR 'life quality':ti,ab,kw OR 'living standards':ti,ab,kw OR 'mental health':ti,ab,kw OR perception:ti,ab,kw OR 'physical health':ti,ab,kw OR satisfaction:ti,ab,kw OR 'quality of life':ti,ab,kw OR qol:ti,ab,kw OR safety:ti,ab,kw OR 'standard of living':ti,ab,kw OR 'value of life':ti,ab,kw OR 'well being':ti,ab,kw) AND [2013-2023]/py | 358 |
| 17.05.2023 11:18 | PubMed | ((activities of daily living[Title/Abstract] OR independence[Title/Abstract] OR life quality[Title/Abstract] OR living standards[Title/Abstract] OR mental health[Title/Abstract] OR perception[Title/Abstract] OR physical health[Title/Abstract] OR satisfaction[Title/Abstract] OR quality of life[Title/Abstract] OR qol[Title/Abstract] OR safety[Title/Abstract] OR standard of living[Title/Abstract] OR value of life[Title/Abstract] OR well-being[Title/Abstract]) OR (activities of daily living OR independence OR life quality OR living standards OR mental health OR perception OR physical health OR satisfaction OR quality of life OR qol OR safety OR standard of living OR value of life OR well-being[MeSH Terms])) AND ((digital assistive tool*[Title/Abstract] OR digital assistive technolog*[Title/Abstract] OR gerontechnolog*[Title/Abstract] OR mobile[Title/Abstract] OR robot*[Title/Abstract] OR supportive technolog*[Title/Abstract] OR technolog* assistive device*[Title/Abstract] OR voice assistant*[Title/Abstract] OR wearable device*[Title/Abstract] OR wearable technolog*[Title/Abstract]) OR (digital assistive tool* OR digital assistive technolog* OR gerontechnolog* OR mobile OR robot* OR supportive technolog* OR technolog* assistive device* OR voice assistant* OR wearable device* OR wearable technolog*[MeSH Terms])) AND (Search: (alzheimer*[Title/Abstract] OR dement*[Title/Abstract] OR early-onset[Title/Abstract] OR frontotemporal lobar degeneration[Title/Abstract] OR lewy-body dementia[Title/Abstract] OR mixed dementias[Title/Abstract] OR vascular dementia[Title/Abstract] OR young onset[Title/Abstract]) OR (alzheimer* OR dement* OR early-onset OR frontotemporal lobar degeneration OR lewy-body dementia OR mixed dementias OR vascular dementia OR young onset[MeSH Terms])), from 2013 - 2023 | 3736 |
| 17.05.2023 11:30 | Scopus | ( TITLE-ABS-KEY ( "alzheimer"  OR  "dement"  OR  "early-onset"  OR  "frontotemporal lobar degeneration"  OR  "lewy-body dementia"  OR  "mixed dementias"  OR  "vascular dementia"  OR  "young onset" )  AND  TITLE-ABS-KEY ( "digital assistive tool"  OR  "digital assistive technology"  OR  "mobile"  OR  "gerontechnology"  OR  "robot"  OR  "supportive technology"  OR  "technology assistive device"  OR  "voice assistant"  OR  "wearable device"  OR  "wearable technology" )  AND  TITLE-ABS-KEY ( "activities of daily living"  OR  "independence"  OR  "life quality"  OR  "living standards"  OR  "mental health"  OR  "perception"  OR  "physical health"  OR  "satisfaction"  OR  "quality of life"  OR  "qol"  OR  "safety"  OR  "standard of living"  OR  "value of life"  OR  "well-being" ) )  AND  ( LIMIT-TO ( PUBYEAR ,  2023 )  OR  LIMIT-TO ( PUBYEAR ,  2022 )  OR  LIMIT-TO ( PUBYEAR ,  2021 )  OR  LIMIT-TO ( PUBYEAR ,  2020 )  OR  LIMIT-TO ( PUBYEAR ,  2019 )  OR  LIMIT-TO ( PUBYEAR ,  2018 )  OR  LIMIT-TO ( PUBYEAR ,  2017 )  OR  LIMIT-TO ( PUBYEAR ,  2016 )  OR  LIMIT-TO ( PUBYEAR ,  2015 )  OR  LIMIT-TO ( PUBYEAR ,  2014 )  OR  LIMIT-TO ( PUBYEAR ,  2013 ) ) | 314 |
| 17.05.2023 11:36 | Web of Science | (alzheimer* OR dement* OR early-onset OR frontotemporal lobar  1 degeneration OR lewy-body dementia OR mixed dementias OR vascular dementia OR young onset (Topic)) AND (digital assistive tool* OR digital assistive technolog* OR gerontechnolog* OR mobile OR robot* OR supportive technolog* OR technolog* assistive device* OR voice assistant* OR wearable device* OR wearable technolog* (Topic)) AND (activities of daily living OR independence OR life quality OR living standards OR mental health OR perception OR physical health OR satisfaction OR quality of life OR qol OR safety OR standard of living OR value of life OR well-being (Topic)) AND (2023 OR 2022 OR 2021 OR 2020 OR 2019 OR 2018 OR 2017 OR 2016 OR 2015 OR 2014 OR 2013 (Publication Years)) | 997 |
