## Supplementary file 2 for "Impact of digital assistive technologies on the quality of life for people with dementia: A scoping review"

Supplementary Table C. Data Charting Form.

| **Category** | **Example / Definition** |
| --- | --- |
| **Title of the study** |  |
| **DOI** |  |
| **Year of publication** |  |
| **Author name/s** |  |
| **Author location/s** |  |
| **Study approach** | i.e., qualitative, quantitative, or mixed-method |
| **Type of article** | i.e., case study, observational study, RCT, review, trial, other… |
| **Study location** | i.e., where the study was carried out |
| **Study Setting** | i.e., domestic environment, long-term care, daycare center, other… |
| **DHT category** | i.e., non-health system software/ digital health solutions, health system operational software, health system clinical software, health & wellness, patient monitoring, care support, digital diagnostics, or digital therapeutics |
| **Explanation of DAT** | i.e., specification of the digital assistive technology (e.g., name/brand) |
| **Target population** |  |
| **Outcome measured** | i.e., the primary outcome being measured in the study |
| **Aim/s of the study** |  |
| **Study methods summary** |  |
| **Key findings** | i.e., study findings relevant to study objectives |
| **QOL measure** | i.e., how is QOL measured (e.g., rating of QOL through a questionnaire, activity instrument, cognitive status, etc.) |
| **Reported effect** | i.e., Yes or No |
| **Notes** |  |
